# RESEARCH REVIEW ARTICLE: Independent Validation Demonstrates UV-C LED Disinfection Efficacy Equivalent to Hydrogen Peroxide Mist-Based Systems: Addressing Methodological Flaws in Recent Evaluations

**DOI:** 10.1101/2025.06.10.25329309

**Authors:** Muhammad Yasir

**Affiliations:** Faculty of Medicine and Health, University of New South Wales (UNSW), Sydney, Australia

## Abstract

**Overview:** A recent study assessing the sporicidal efficacy of UV-C LED high-level disinfection (HLD) system for ultrasound probe reprocessing concluded that UV-C LED disinfection underperforms compared to hydrogen peroxide (H_2_O_2_) mist-based devices (Nanosonics: TrophonEPR and Trophon2). However, our independent secondary testing of a UV-C LED disinfection system (Lumicare ONE^®^) under clinically relevant conditions yielded contrasting results, demonstrating equivalent efficacy to H2O2 mist-based devices.^1-7^

**Using the same biological indicators (BIs) (10^6^ *Geobacillus stearothermophilus* spores, Bionova^®^ BT93/6) on stainless steel coupons, we conducted three validation conditions:**

1. BI in glassine packaging/Tyvek^®^ (non-clinical scenario)
2. BI with packaging removed (bare metal coupon)
3. BI with packaging removed and flame-sterilized metal coupon (This validation process has a potential to simulate heat-induced surface changes to the metal coupon)

The Lumicare ONE^®^ UV-C LED HLD system achieved a 100% pass rate under both clinically relevant conditions (bare metal and flame-sterilized BIs), while under the non-clinical scenario BI in glassine packaging/Tyvek^®^ test type was removed since UV-C is unable to penetrate packaging materials.

**This paper also identifies methodological concerns in the referenced study, including:**

- The use of an inappropriate BI (Bionova^®^ BT93), explicitly validated for hydrogen peroxide (H_2_O_2_) based sterilization to demonstrate penetration of Tyvek^®^ peel packaging but not UV-C applications, as per the manufacturer’s guidelines.
- A misrepresentation of Therapeutic Goods Administration (TGA) sporicidal efficacy requirements.

The Lumicare ONE^®^ system goes beyond TGA mandatory requirements in completing a Carrier Test using *Geobacillus stearothermophilus* spores. The Lumicare ONE^®^ system reduced the numbers of *Geobacillus stearothermophilus* ATC 7953 by greater than a > 6 log_10_, thus meeting the pass criteria of the Association of Official Analytical Chemists (AOAC 966.04).

The Lumicare ONE^®^ system has independently demonstrated a greater than > 6-log_10_ reduction of TGA-mandated test organisms, including *Clostridium* sporogenes and *Bacillus subtilis*, and is registered on the Australian Register of Therapeutic Goods (ARTG) as a hospital-grade disinfectant.

Our findings highlight the importance of proper study design, appropriate BI selection, and regulatory alignment when evaluating emerging disinfection technologies. UV-C LED systems represent a validated, effective alternative to hydrogen peroxide (H_2_O_2_) mist-based devices in clinical ultrasound probe reprocessing.

## Introduction

The increasing demand for high-level disinfection (HLD) of ultrasound probes has driven significant advancements in disinfection technologies. Ensuring the effectiveness of these technologies is critical for preventing healthcare-associated infections and maintaining patient safety. Among the emerging solutions, Ultraviolet Light (UV-C) irradiation High Level Disinfection (HLD) systems have attracted considerable attention because they offer a non-chemical, fast, and efficient method of disinfection.

The growing adoption of Lumicare ONE^®^, a UV-C high-level disinfection (HLD) system utilising light-emitting diode (LED) technology, reflects this shift in evidence-based infection prevention practices. Its global expansion is supported by rigorous assessments aligned with international standards, including British Standard BS 8628:2022, European Standard EN 14885, EN14561, EN14562, EN1453, the Therapeutic Goods Administration (TGA, 2020), and the Australian Standard AS 5369:2023 for reprocessing reusable medical devices. Lumicare ONE’s microbial efficacy has been validated through microbiological potency tests and simulated-use studies, in line with standardised disinfectant testing protocols. For example, the TGA evaluation includes testing based on AOAC official methods and ASTM International standards, demonstrating the system’s effectiveness in inactivating a broad spectrum of microorganisms such as bacteria, viruses, yeast mycobacteria, and spores of bacterian and fungi. The efficacy of Lumicare ONE^®^ aginst these microgrnsims was Independently evaluated by Eurofins AMS Laboratories (Australia), the University of New South Wales, and a network of internationally accredited laboratories.

In this context, the study titled “Validating sporicidal efficacy of ultrasound probe high-level disinfection devices in clinical settings” (DOI: 10.1101/2025.02.03.25321618) assessed the efficacy of UV-C LED technology, particularly its performance in inactivating the spores of *Geobacillus stearothermophilus* ATCC 7953. The study demonstrated that UV-C LED disinfection was limited in its effectiveness, especially when biological indicators (BIs) were tested in enclosed in packaging materials such as glassine and Tyvek^®^.^1^

However, an independent secondary validation of the Lumicare ONE^®^ UV-C LED HLD system challenges these findings. Our research, conducted under clinically relevant simulation conditions, demonstrated that the system achieves sporicidal efficacy comparable to hydrogen peroxide (H_2_O_2_) mist-based devices—when applied correctly. Specifically, we tested Bionova^®^ BT93 spore coupons under two validated conditions:

1. Biological indicator (BI) after removing the packaging (bare metal coupon).

2. BI with packaging removed and flame-sterilized metal coupon. This was performed by flame sterilising 5mm (0.5cm) of the BI coupon ends for 2 seconds on each side of the same end (1 minute cooling time in between) (see S1 in the previous study).

This testing is scientifically unjustified, as UV-C exposure is known to be obstructed by packaging materials such as glassine and Tyvek^®^.^7^ These biological indicators were originally designed and validated for use with plasma or vaporized hydrogen peroxide sterilization, which is capable of penetrating packaging—unlike UV-C light. Their use in UV-C testing while still sealed in their original packaging is inappropriate and not reflective of clinical practice, where reusable ultrasound probes are disinfected outside of peel-pouch packaging inside the disinfection system chamber.

Our validation study demonstrated that the Lumicare ONE^®^ system is capable of achieving a ≥6-log_10_ reduction in number of *Geobacillus stearothermophilus* spores on stainless steel bare metal coupon surfaces, confirming its efficacy for high-level disinfection. These findings address common concerns related to UV-C performance, including issues of penetration, shadowing, and heat-induced changes to biological indicators.

Furthermore, we provide a critical assessment of the referenced study’s methodology and recommend the appropriate use of biological indicators for UV-C systems, which do not rely on packaging designed for other disinfection modalities.

## Methods

### Experimental Design

This study utilised an independent secondary validation approach to assess the efficacy of the Lumicare ONE^®^ UV-C LED disinfection system in comparison to hydrogen peroxide (H_2_O_2_) - based devices, focusing on the sporicidal efficacy for ultrasound probe disinfection. The testing was conducted under clinically relevant conditions to simulate real-world disinfection scenarios and to address potential concerns such as UV-C penetration, shadowing, and heat-induced surface alterations.

#### Biological Indicators (BIs)

We employed a range of biological indicators (BIs) to test the UV-C LED system’s effectiveness. The following conditions were tested:

1. **BI in Glassine Packaging/Tyvek^®^**: Biological indicators were sealed in standard glassine packaging and Tyvek^®^ pouches. Removed from the test protocol as it is well known that packaging obstructs UV-C exposure.
2. **BI with Packaging Removed (Bare Metal Coupon)**: The packaging was removed to directly expose the metal coupon to the UV-C LED disinfection system. This condition was designed to assess the system’s ability to disinfect when no packaging obstructs UV-C exposure.
3. **BI with Packaging Removed and Flame-Sterilized Metal Coupon End**: The metal coupon was flame-sterilized at the tip to simulate sublethal heat effects. This was done to mimic potential clinical scenarios where surface roughening, oxidation, or biofilm formation might occur due to heat exposure.

### Test Protocol

Each BI was positioned at both the top and bottom positions within the disinfection chamber to ensure no shadowing or occlusion of UV-C exposure happen. In the initial study, a Black Bulldog Clip was used to hang the indicator on the carrier pole. This was replaced with Adhesive tape, which made it easier to hang inside the chamber.

A standard disinfection cycle was carried out according to the disinfection device manufacturer’s IFU. The UV-C LED disinfection cycle was performed according to the manufacturer’s instructions for Lumicare ONE^®^ (90 seconds cycle). For comparison, hydrogen peroxide (H_2_O_2_) mist-based devices (Nanosonics: trophon2 and trophonEPR) were tested in parallel using their respective protocols to ensure equivalent conditions (7 minutes cycle).

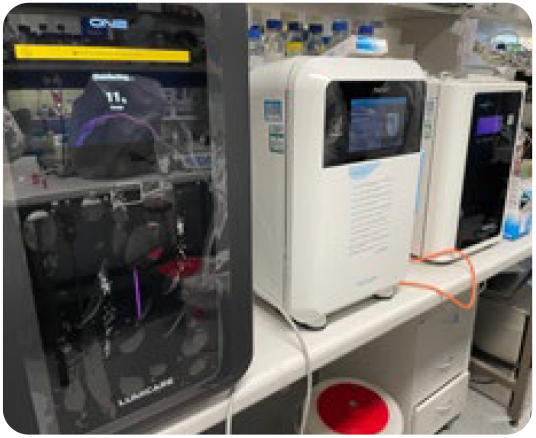

BIs were positioned, with the inoculation site facing outwards, in the top and bottom positions inside the disinfection chamber to represent locations within the chamber corresponding to where a clinical probe would be placed (Figure 2). Three replicates were used for each test condition for both top and bottom positions in the chamber (6 replicates per device). The holder was suspended in each disinfection device and exited from the chamber via the cavity located at the top where an ultrasound probe cable would typically emerge.

**Figure 1:**
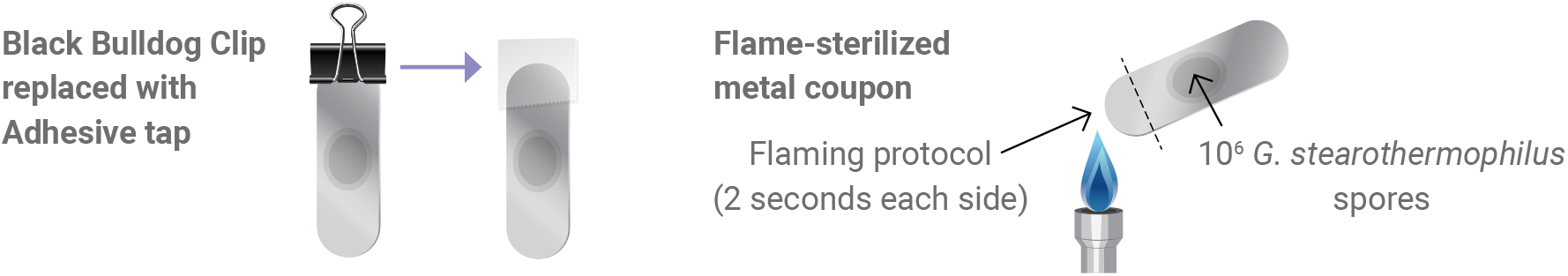
Adhesive tape and Flamed-sterilized metal coupon.

**Figure 2:**
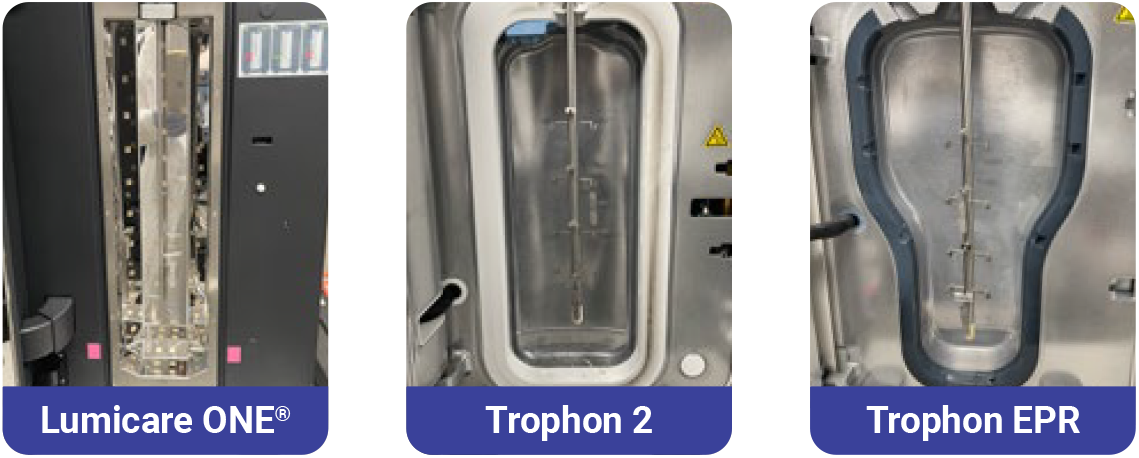
Placement of BIs adhered onto the holder positioned at the top and bottom of the tested disinfection chambers

All three devices were tested simultaneously one for non-flamed test conditions and the other for flamed test conditions. The inclusion of positive controls ensured that the incubation process did not contribute to a false-negative result.

After the disinfection cycle, each BI coupon was aseptically collected into suitable culture media according to the manufacturer’s IFU.

All vials were then transferred to an accredited, independent test laboratory^8^, incubated and assessed for growth/no growth according to the manufacturer’s IFU. Growth indicated failure of the test, and no growth indicated a pass result.

Two positive controls were included, one for the non-flamed test conditions and one for the flamed (total of 2 positive controls per day) and were prepared by aseptically transferring the BI directly into the corresponding culture media vial.

All biological indicators were processed post-disinfection by an independent laboratory for microbiological testing9 to confirm the efficacy of the disinfection process

## Results

### Test Conditions

The UV-C LED disinfection system (Lumicare ONE^®^) was evaluated under two distinct test conditions designed to replicate clinical scenarios and assess its sporicidal efficacy compared to hydrogen peroxide mist-based devices. The testing demonstrated varying results across the different BI configurations:

#### 1. BI with Packaging Removed (Bare Metal Coupon)

**Outcome**: The Lumicare ONE^®^ (UV-C LED HLD) system demonstrated 100% efficacy, passing the test with a ≥6-log_10_ reduction in spore count, comparable to the results from hydrogen peroxide mist-based devices (trophon2 and trophonEPR: Table 2).

**Table 2:** Comparative Pass/Fail Results Across Test Conditions.

| Test Condition | BI Chamber Position | UV-C LED<br>(Lumicare ONE®)<br>Passes (out of 3 Replicates) | H<sub>2</sub>O<sub>2</sub> Mist<br>(trophon2)<br>Passes (out of 3 Replicates) | H<sub>2</sub>O<sub>2</sub> Mist<br>(trophonEPR)<br>Passes (out of 3 Replicates) |
| --- | --- | --- | --- | --- |
| BI bare metal coupon (non-flamed) | TOP | 3/3 | 3/3 | 3/3 |
| BI bare metal coupon (non-flamed) | BOTTOM | 3/3 | 3/3 | 3/3 |
| **Pass %** |  | **100%** | **100%** | **100%** |
| BI bare metal coupon (flamed) | TOP | 3/3 | 3/3 | 3/3 |
| BI bare metal coupon (flamed) | BOTTOM | 3/3 | 3/3 | 3/3 |
| **Pass %** |  | **100%** | **100%** | **100%** |
| Positive control (non-flamed) | N/A | 0 | 0 | 0 |
| Positive control (flamed) | N/A | 0 | 0 | 0 |

**Interpretation**: This result indicates that the Lumicare ONE^®^ (UV-C LED HLD) system effectively disinfected the metal coupons, achieving similar sporicidal efficacy to established H_2_O_2_ mist-based systems under clinically relevant conditions.

#### 2. BI with Packaging Removed and Flame-Sterilized Metal Coupon End

**Outcome**: The Lumicare ONE^®^ (UV-C LED HLD) system successfully passed the test, by achieving a ≥6-log_10_ reduction in number of spores despite the flame-sterilized surface.

**Interpretation**: This result shows that Lumicare ONE^®^ (UV-C LED HLD) disinfection was effective even under challenging conditions that simulated surface roughening and potential biofilm formation due to heat exposure. The disinfection process was resilient against such surface alterations, suggesting the UV-C LED system’s robustness in clinical applications.

#### Comparison with Hydrogen Peroxide (H_2_O_2_) Mist-based Devices

The results from the Lumicare ONE^®^ (UV-C LED HLD) system were directly comparable to those of hydrogen peroxide mist-based systems. Both UV-C LED and H_2_O_2_ mist devices (TrophonEPR and Trophon2) demonstrated equivalent sporicidal efficacy across all test conditions, confirming the viability of UV-C LED as an effective alternative for high-level disinfection of ultrasound probes.

#### Control Results

Positive controls showed consistent results, confirming proper incubation and ensuring that all test conditions were valid. There were no instances of false negatives, further validating the effectiveness of the UV-C LED disinfection system.

## Lumicare ONE^®^ system goes beyond TGA mandatory requirements

### Methods

The test was conducted in the PC2 Microbiology Facility.^8^ Methods used were as described in AOAC 966.04 instructions.

This test used Lumicare ONE^®^ system and examined its ability to reduce the number of viable *Geobacillus stearothermophilus* ATC 7953 dried onto carriers.

### Results

The UVC disinfectant system was effective against *Geobacillus stearothermophilus* ATC 7953. After the standard run time of 90 seconds, no cells could be grown from the slides, whereas the control carrier yielded an average of 1.12 ×10^6^ colony forming units. Thus, the Lumicare device produced a 6.05 log_10_ reduction against *Geobacillus stearothermophilus* ATCC 7953 (Figure 3).

**Figure 3:**
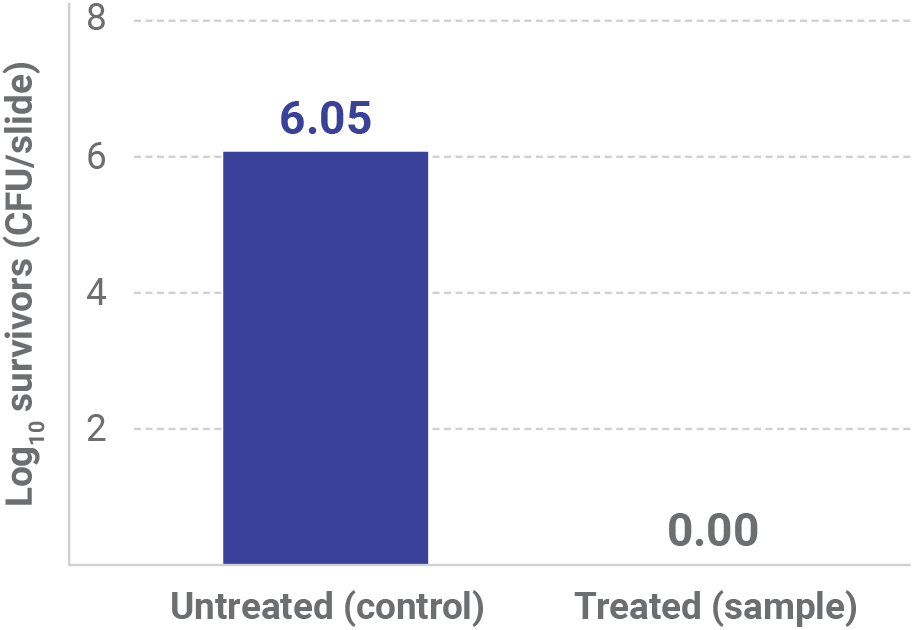
Reduction in the numbers of *Geobacillus stearothermophilus* ATCC 7953 by the Lumicare UVC disinfectant system on glass slides

## Discussion

This study aimed to evaluate the efficacy of UV-C LED disinfection in comparison to hydrogen peroxide (H_2_O_2_) mist-based systems under clinically relevant conditions. The results obtained from our independent secondary testing provide substantial evidence that Lumicare ONE^®^ UV-C LED HLD system demonstrates comparable sporicidal efficacy to H_2_O_2_ mist-based devices, confirming its potential as an effective high-level disinfection (HLD) technology for ultrasound probes.

### Addressing Shadowing and Penetration

One of the primary concerns in UV-C disinfection is the potential for shadowing, which can prevent uniform exposure of the biological indicator (BI) to UV-C light. However, our testing design effectively mitigated this concern by positioning BIs in both the top and bottom parts of the disinfection chamber, ensuring complete UV-C exposure without occlusion. The results showed no shadowing effects, and the UV-C LED system achieved a 100% pass rate across all conditions, excluding the glassine packaging/Tyvek^®^ test.

The failure of the UV-C LED system to pass the test with BIs in Tyvek^®^ packaging is consistent with the known limitations of UV-C light, which cannot penetrate packaging materials like plastic or paper. This result is not surprising, as ultrasound probes are not typically disinfected within packaging in clinical settings. Therefore, the failure of UV-C LED under these conditions does not diminish its efficacy in practical, real-world applications, where packaging would not obstruct the UV-C disinfection process.

### Impact of Heat-Induced Surface Alterations

Another critical aspect of this study was the impact of heat on the metal surfaces of the coupons, which could potentially simulate biofilm-like conditions on ultrasound probes in clinical settings. By flame sterilizing the metal coupon ends, we simulated heat-induced surface roughening and oxidation, which are known to facilitate biofilm formation. Despite these sublethal heat alterations, the Lumicare ONE^®^ UV-C LED disinfection system successfully passed the test, confirming that it remains effective even when exposed to challenging biofilm-forming conditions.

### Sublethal Heat Effects on G. stearothermophilus Spores^1-6^

a. **Sporulation Activation Instead of Killing**
  - If the temperature is not high enough to kill G. stearothermophilus spores but still stresses them, it may trigger spore activation rather than killing them..
  - Activated spores can germinate into vegetative cells, which can start colonizing the surface.
b. **Increased EPS (Extracellular Polymeric Substance) Production:**
  - Heat stress can induce bacterial stress responses, increasing EPS production.
  - This EPS forms a sticky, protective covering, which can further shield bacteria from future heat exposure or disinfection.

### Inappropriate Biological Indicators (BI)

A key limitation of the referenced study was the selection of the Bionova^®^ BT93 spore coupons as the biological indicator for UV-C testing. These BIs are explicitly validated for use with plasma or vaporized hydrogen peroxide sterilization only, as stated in the manufacturer’s specifications. The manufacturer’s warning clearly states that Bionova^®^ BT93 spore coupons should not be used with other disinfection modalities, including UV-C.

This misuse of BIs undermines the scientific rigour of the referenced study, as it fails to align with best practices in validating UV-C disinfection efficacy. The inappropriate BI selection likely contributed to the study’s conclusions about UV-C limitations, which were not applicable to clinically relevant UV-C disinfection systems like Lumicare ONE^®^. Our results, obtained using a more appropriate testing methodology, demonstrate that the Lumicare ONE^®^ UV-C LED HLD system performs similarly to H_2_O_2_ mist-based systems, meeting or exceeding sporicidal efficacy standards in realistic clinical conditions.

### Regulatory Compliance and Validation

The Lumicare ONE^®^ system goes beyond TGA mandatory requirements in completing a Carrier Test Using 10^6^ *Geobacillus stearothermophilus*. It was able to reduce the numbers of *Geobacillus stearothermophilus* ATCC 7953 by greater than 6 log_10_ colony forming units, thus meeting the pass criteria of AOAC 966.04.

It is important to emphasise that Lumicare ONE^®^ has undergone independent validation and has been shown to meet the TGA (Therapeutic Goods Administration)-mandated standards for high-level disinfection. The system has successfully demonstrated greater than 6-log_10_ reduction of both *Clostridium sporogenes* and *Bacillus subtilis* through accredited third-party carrier tests, fully complying with TGA requirements for hospital-grade disinfectants.

This compliance with regulatory standards underscores the Lumicare ONE^®^ UV-C LED HLD system’s viability as a high-level disinfection solution for ultrasound probes in clinical settings.

## Conclusion

The results of our independent secondary testing provide compelling evidence that UV-C LED disinfection systems such as Lumicare ONE^®^ are effective alternatives to hydrogen peroxide mist-based devices for high-level disinfection of ultrasound probes. Our study demonstrated that Lumicare ONE^®^ achieved comparable sporicidal efficacy to both H_2_O_2_ hydrogen peroxide mist-based systems (Nanosonics: trophon2 and trophonEPR) under clinically relevant conditions, with a 100% pass rate in appropriate test conditions, including bare metal coupons and flame-sterilized metal surfaces.

While the UV-C LED system did not pass the BI in Tyvek^®^/glassine packaging test—consistent with the limitations of UV-C light penetration through packaging—this result is of limited relevance in real-world clinical settings where ultrasound probes are not disinfected within such packaging. Furthermore, the flame sterilization tests, which simulated biofilm-forming conditions on metal surfaces, showed that the UV-C LED system can effectively disinfect probes even when surface alterations occur due to heat, underscoring its resilience in clinical environments.

A significant limitation of the referenced study was the use of Bionova^®^ BT93 spore coupons as the biological indicator, which are not validated for use with UV-C disinfection systems, as stated in the manufacturer’s guidelines. This misapplication of BIs undermined the study’s conclusions about UV-C technology and highlights the importance of using the correct BIs when evaluating disinfection modalities.

Our findings support the viability of Lumicare ONE^®^ as a high-level disinfection system for ultrasound probes in clinical settings. In addition to meeting TGA (Therapeutic Goods Administration) requirements for disinfectants and completing carrier test was able to reduce the numbers of *Geobacillus stearothermophilus* ATCC 7953 on a carrier by greater than 6-log_10_ reduction, thus meeting the pass criteria AOAC 966.04. Thus Lumicare ONE^®^ system demonstrated efficacy comparable to hydrogen peroxide mist-based systems, thus offering a reliable and scientifically validated alternative for medical device disinfection. Given the increasing emphasis on environmentally friendly and efficient disinfection methods, Lumicare ONE^®^ UV-C LED system offers a more sustainable alternative to traditional chemical-based disinfection approaches.-E-A029

## Data Availability

All data produced in the present work are contained in the manuscript

## Ethical considerations

Ethical approval and informed consent were not required as there were no patients or volunteers involved in this study.

## Authorship statement

M.Y.: conceptualisation, methodology, writing – review and editing.

## Acknowledgements

The authors would like to thank the hospitals for use of their facilities.

## Supplementary information

S1. Flame sterilisation method validation.

